# Dynamic magnetic resonance imaging of muscle contraction in facioscapulohumeral muscular dystrophy

**DOI:** 10.1101/2021.05.22.21257219

**Authors:** Xeni Deligianni, Francesco Santini, Matteo Paoletti, Francesca Solazzo, Niels Bergsland, Giovanni Savini, Arianna Faggioli, Giancarlo Germani, Mauro Monforte, Enzo Ricci, Giorgio Tasca, Anna Pichiecchio

**Affiliations:** Radiology/ Division of Radiological Physics, University Hospital of Basel, Basel, Switzerland; Biomedical Engineering, University of Basel, Allschwil, Switzerland; Advanced Imaging and Radiomics Center, Neuroradiology Department, IRCCS Mondino Foundation, Pavia, Italy; School of Specialization in Clinical Pharmacology and Toxicology Center of research in Medical Pharmacology, School of medicine University of Insubria, Varese, Italy; Buffalo Neuroimaging Analysis Center, Department of Neurology, Buffalo Neuroimaging Analysis Center, Department of Neurology, Jacobs School of Medicine and Biomedical Sciences, Buffalo, NY, United States; IRCCS Fondazione Don Carlo Gnocchi ONLUS, Milan, Italy; Unità Operativa Complessa di Neurologia, Fondazione Policlinico Universitario A. Gemelli IRCCS, Rome, Italy; Department of Brain and Behavioral Sciences, University of Pavia, Pavia, Italy

**Keywords:** muscle MRI, neuromuscular electrical stimulation, FSHD, dynamic MRI, strain, muscular dystrophy

## Abstract

**Background:** Quantitative muscle MRI (water-T2 and fat mapping) is being increasingly used to assess disease involvement in muscle disorders, while imaging techniques for assessment of the dynamic and elastic muscle properties have not been translated yet into clinics.

**Methods:** Here, we quantitatively characterized muscle deformation (strain) in patients affected by facioscapulohumeral muscular dystrophy (FSHD), a prevalent muscular dystrophy, by applying dynamic MRI synchronized with neuromuscular electrical stimulation (NMES). We evaluated the quadriceps muscles in 34 ambulatory patients and 12 healthy controls, at 6 month intervals.

**Results:** We found that while a subgroup of patients behaved similarly to controls, for another subgroup the strain significantly decreased over time (50% over 1.5 years). Dynamic MRI parameters did not correlate with quantitative MRI.

**Conclusions:** In conclusion, our results suggest that the evaluation of muscle ability to contract by NMES-MRI could be used to explore the elastic properties and monitor muscle involvement in FSHD and other neuromuscular disorders.

## BACKGROUND

Facioscapulohumeral muscular dystrophy (FSHD) is one of the most prevalent muscular dystrophies (1,2), which are genetic disorders characterized by progressive degeneration of the skeletal muscle. In muscular dystrophies, the muscle undergoes several pathophysiological processes encompassing necrosis, inflammation, fibrosis, and finally replacement by adipose tissue. The progression of muscle wasting and weakness in FSHD is particularly stepwise (3,4) and asymmetric (5,6) although generally slow. The clinical phenotype of the disease can overlap with other types of muscle disorders (7), and magnetic resonance imaging (MRI) can identify patterns of involvement that help the differential diagnosis (3).

Therefore, MRI in the last years has been increasingly applied to evaluate the extent of muscle involvement and evolution in FSHD (3,8–10). Overall, there is strong evidence pointing to the direction that hyperintensities on short tau inversion recovery (STIR) images reflect an active phase (3), which is followed by fat infiltration (11) typically identified on T1 weighted (T1w) images, as the disease progresses. Indeed, the results of several studies suggested that muscles presenting with STIR hyperintensity have a faster progression towards fat replacement (5,12).

Functional measurements such as the 6-minute walking test (6MWT) (13) or force measurements (14) are also relevant in the follow-up of neuromuscular patients, and for FSHD as well. In a more recent study, it was observed that changes of fat fraction in FSHD occur more at a single muscle level, and the larger these changes are, the faster the change in muscle strength (15).

Overall, while different MRI techniques that are available to date are able to identify muscle necrosis/inflammation (taking advantage mainly of T2-weighted sequences) and fat replacement (with use of fat fraction measurements) with very good accuracy, in certain cases they may not be sufficiently sensitive in muscular dystrophies (16). In addition, the deposition of collagen tissue, accounting for fibrosis and supposed to affect the elastic properties of skeletal muscle, is currently escaping detection by MRI. In the specific case of FSHD, given the slow progression of the disease, even fat fraction changes might be hard to detect (15) and, as a consequence, capturing disease progression in the short timeframe of a clinical trial can be challenging.

Muscle contractility is another useful quantitative parameter in muscular dystrophies, as available literature on ultrasound-based methods suggests (17–19). For example, in combination with MRI, it can help to guide clinical decisions in Duchenne muscular dystrophy patients (20). Nonetheless, the application of an MRI-based method has various advantages over the ultrasound technique, such as lower dependency on operator experience and the potential for increased volumetric coverage of the muscle and three-dimensional visualization.

Neuromuscular electrical stimulation (NMES)-synchronized MRI of muscle deformation has been used to characterize how muscles, and in particular the more superficial ones, contract (21–23). This method allows to estimate the contraction velocity as well as to characterize the deformation of muscle with strain maps. The strain is defined as the change of length per unit length in each spatial direction of a material under stress, in this case the muscle, with respect to its length at rest. In a previous study using NMES-synchronized MRI, the strain from phase-contrast MRI images has been shown to be different between ten young and ten senior (>70 y old) healthy volunteers (23). Moreover, the rates at which the strain reaches the maximum (*buildup constant*)(23) and relaxes to zero (*release constant)* can offer additional information about the ability of the muscle to deform. A healthier muscle is expected to have a faster build-up of strain, higher maximum, and faster release as well in electromyography-based assessments (24). However, to our knowledge, there is no study to date on the application of similar dynamic MRI methods, either with evoked NMES or voluntary contraction, in patients with neuromuscular diseases.

The purpose of this study was to characterize muscle deformation behavior in FSHD patients compared to healthy controls followed longitudinally. For this aim, NMES was applied for the periodic contraction of the quadriceps muscle in synchronization with phase-contrast MRI to acquire dynamic data. Finally, the results from the dynamic analysis were compared to water T2 relaxation and fat fraction measurements.

## METHODS

### Study design & subjects

34 ambulatory patients were included in this investigation, enrolled in a larger study on longitudinal MRI biomarkers in FSHD and with a confirmed genetic diagnosis. Patients were clinically evaluated at baseline using the Clinical Severity Score (25), a specific score for FSHD ranging from 0 (asymptomatic patient) to 5 (wheelchair-bound patient), the 6MWT, which assesses the maximum distance covered during a 6 minutes-walk, and the dynamometric evaluation (microFET® handheld dynamometer [Hoggan Scientific, Salt Lake City, UT]) of the maximum voluntary contraction of the quadriceps. For all patients, the length of the 4q35 BlnI resistant, p13-E11 EcoRI fragment causing the disease was reported (in kb).

The patients (13 female, 21 male) at the time point of the first scan were 44.8 ± 9.0 years old with weight 77.5 ± 20.4 kg and height 173.1 ± 9.3 cm (i.e., average weight and height of all datasets over all 3 time points).

The healthy controls (HCs) (8 female, 4 male), chosen mainly between non-consanguineous family members of FSHD subjects and with no known neurological disease were 49.9 ± 12.2 years old with weight 70.5 ± 9.5 kg and height 168.2 ± 7.8 cm.

All patients were recruited for a comprehensive quantitative muscle MRI protocol and a dynamic MRI scan at four time points six months apart (t0, t1, t2, t3), whereas HCs were scanned up to three times. Dynamic scans were performed for both thighs separately, so two dynamic datasets were ideally collected for each participant. The study was approved by the local institutional review boards (at the Mondino Foundation by the “Comitato Etico Area Referente Pavia Fondazione IRCCS Policlinico San Matteo” - Prot. n. P-20200042370 and the Policlinico Gemelli’s Ethics Committee - Prot. n. 7451/18 ID 1952) and all volunteers signed an informed participation agreement.

### Imaging & Stimulation Protocol

For the dynamic MRI acquisition, a commercial NMES device (InTENSity Twin Stim III TENS and EMS Combo [Current Solutions LLC, Austin, TX]) was synchronized with the MRI acquisition. 5.1 x 8.9 cm^2^ rectangular self-adhesive gel-based NMES electrodes [TensUnits.com, Largo, FL]) were employed for stimulation and exact measures of their position were taken for placing them accurately in subsequent visits (i.e., distance between electrodes, distance from patella to the electrodes, lateral position on MRI localizer images, see Fig. 1). The median distance of the electrodes for both legs for FSHD patients was 11.0 cm (1st Quartile, 3rd Quartile=10,14) and for HCs 10.5 (1st Quartile, 3rd Quartile=10.0,12.25). A glycerol marker was positioned in the middle of each of the four electrodes for easier localization on the MRI images. The stimulator electrodes were placed on both sides prior to the scan by identifying the motor point with a stimulation pen (26), and the current was set to a sufficient level to evoke muscle twitching without knee extension (i.e., isometric contraction) and without discomfort or local pain reported from the patient. A training NMES session was performed outside of the scanner room to familiarize the volunteers with the procedure.

**Figure 1.**
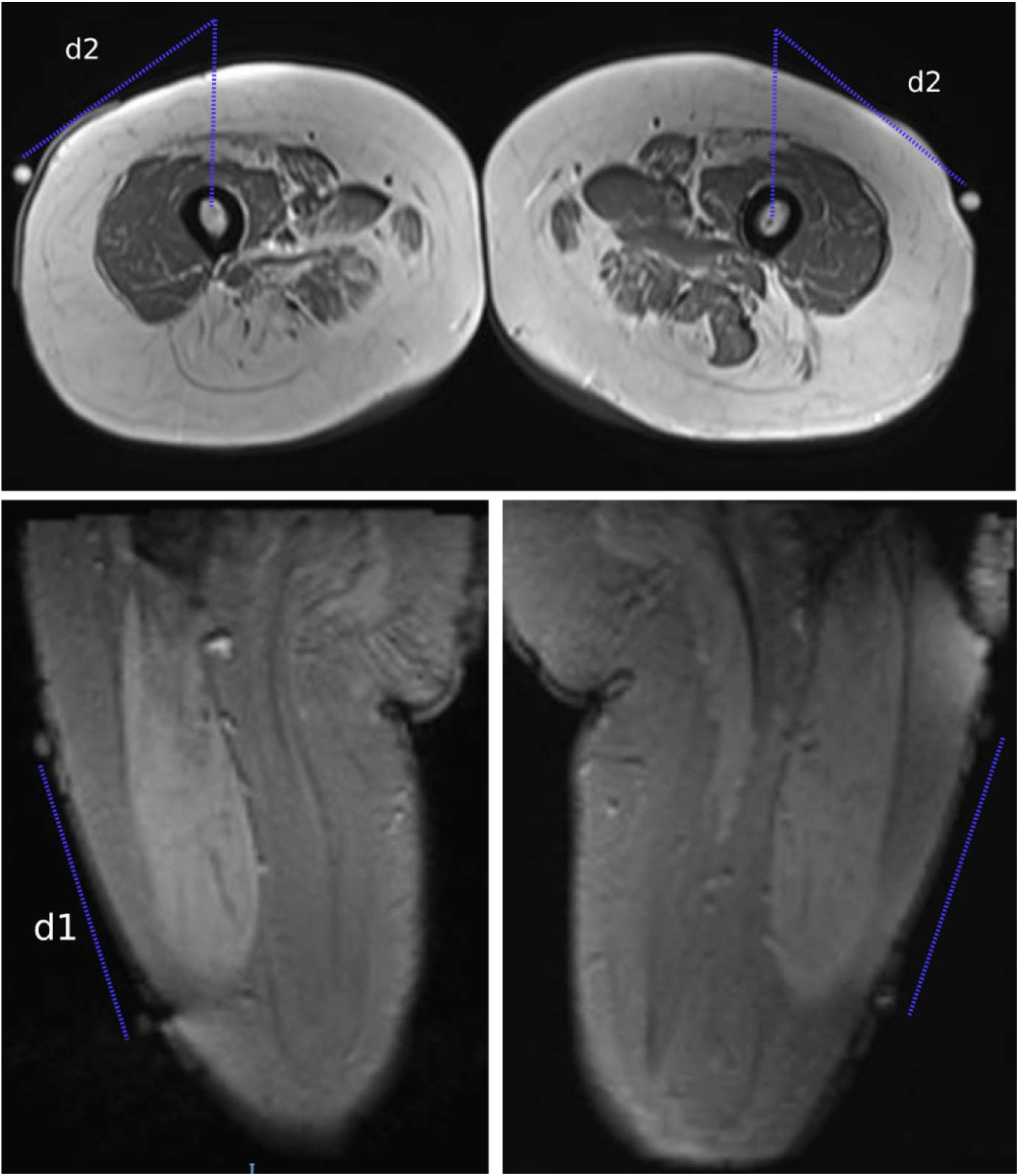
Description of positioning and repositioning of the electrodes. Two electrodes were positioned on each thigh. A glycerol pill was placed on each electrode for easier visualization on the MR images. The second one was positioned after motor point identification before entering the scanner room. The distance between the electrodes (d1), the distance of the glycerol marker to the plane of the bone as measured on the transverse MRI image (d2) and the distance of the marker from the patella.

For every subsequent scan at a later time point, the current was set at the same level of the previous examination or higher, provided it evoked a similar visible muscle contraction (23). When the volunteer reported discomfort or local pain even applying the same current previously used, the current was lowered while still maintaining the ability to provoke muscle twitching.

All subjects were scanned on a 3T clinical MRI scanner (MAGNETOM Skyra [Siemens Healthineers, Erlangen, Germany]). An 18-element body-array was used, centered on the thigh. Regarding the MRI protocol, a three-directional single-slice cine phase-contrast (PC) velocity encoding acquisition was used in triggered mode during periodic contractions (21,23). The NMES device was set to “synchronous mode,” where its two channels are active and deliver stimulation at the same time. One channel was applied through two electrodes to the subject’s skin for stimulation, while the other was fed as input to a custom-made circuit and used to generate the trigger signal for the MRI data acquisition (21,23). The dynamic scans were performed at the end of all the scanning protocol. A parasagittal slice was acquired with voxels of 2.3×2.3×5 mm^3^ and a temporal resolution of 42 ms. The velocity encoding was 25 cm/s. The parameters for the PC acquisition were the following: repetition time/ echo time (TR/TE)=10.6/ 7.21 ms, bandwidth/ pixel = 400 Hz/Px, flip angle=10°, FOV (field-of-view)=225×300 mm^2^, 1 k-space line per segment, acquisition time 5 min) and 94 temporal phases were acquired. Each contraction cycle lasted 5 s (1 s ramp-up, 1 s contraction, 1 s ramp-down, 2 s relaxation).

The dynamic scans were organized in parallel to a more comprehensive quantitative protocol, the full description of which is beyond the scope of this article (27). Here, for a subgroup of volunteers, 56 dynamic single side datasets (ssd) were compared to the average fat fraction and T2 relaxation values of the entire single muscles of the quadriceps (vastus lateralis (VL), vastus intermedius (VI)). For T2 relaxation mapping, a conventional 2D multi-echo spin-echo (MESE) acquisition protocol was used with the following acquisition parameters: number of echoes 8-17, TR 4100.0 ms, first TE and echo spacing 10.9 ms, bandwidth 250 Hz/px, matrix size 192×384×1, resolution 1.2×1.2×10.0 mm^3^. Then, water T2 fitting was performed with the extended-phase-graph fitting method (28,29). For fat fraction quantification, a multi-echo gradient-echo (MEGE) custom sequence was used with interleaved monopolar echoes and a dedicated reconstruction pipeline (29,30). The parameters of the protocol were the following: number of echoes 6, TR 35.0 ms, first TE/echo spacing 1.7/1.5 ms (interleaved echo sampling-weakly asymmetric sampling), flip angle 7°, bandwidth 1050 Hz/px, in-plane matrix size 396×432, resolution 1.0×1.0×5.0 mm^3^. The acquired images were post-processed with the publicly-available post-processing algorithm FattyRiot (31) to obtain the fat fraction maps. Single-muscle regions of interest (ROIs) were subsequently segmented on the quadriceps by an experienced radiologist. Average water T2 and fat fractions (FFs) were calculated over the whole VL and VI muscles respectively.

### Post-processing

The velocity images were elaborated offline with Matlab (R2019b [The Mathworks, Inc., Natick, MA]). Velocity images were corrected for phase shading, the displacement was calculated with forward/backward integration, and the Langrangian strain was computed (21). The analysis was performed with the assumption that the pixels of the acquired slice do not move out of the slice during the acquisition. The tensors were diagonalized and the positive eigenvalue was considered as the pixel-wise principal strain. The principal strain maps were calculated from the displacement maps and then visualized as described before (21,22). For displacement derivative calculation the MaxPol routine was used for smoother numerical differentiation (22,32,33).

For strain calculation, a ROI was drawn on the magnitude image of the PC acquisition including both VL and VI. As a characteristic value of the eigenvalues of strain, the spatial average over the ROI was calculated for every time frame of the reconstructed single contraction period and the absolute value maximum of this time curve was considered as peak strain. In addition to the maximum principal strain, the difference in respect to the first scan of each subject was calculated (i.e., the difference in strain, as well as the difference in the applied current).

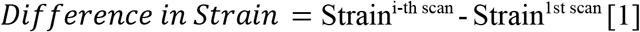

The slope of linear regression between changes in strain and respective changes in stimulation current was used for normalization.

Finally, the time constants at which the strain reached the maximum (*buildup constant*) and relaxed to zero (*release constant*) were also calculated by fitting a sigmoid curve to the corresponding portions of the strain curve (23). The rates were firstly calculated as maps and then the mean values of the respective ROIs were estimated as well as an estimation of error.

In this study, the sigmoid fit is defined as:

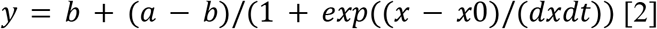

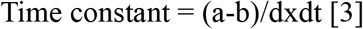

### Statistical Analysis

All graphical plots, linear regression fitting, and correlation coefficient calculations were performed with R/RStudio (34). Specifically, the repeated measures correlation coefficients were calculated, corrected for dependence of datasets that belonged to the same subject (*rmcorr*-function in R) and, where needed, significance levels were corrected with the respective Bonferroni correction. Boxplots were partially replaced by scatter plots, if less than 10 data points were available.

The groups of patients and HCs were tested for differences in age at baseline, body mass index (BMI), weight, height and distance between the electrodes. The values were ln-transformed to compensate partly for skewness and a two-sided t-test was performed acknowledging not equal variance and not paired data. Significance levels were corrected with the respective Bonferroni correction. These comparisons were performed for all datasets at baseline, at time point 1, and for the datasets for which an equal number of data were available at baseline and at time point 1. Finally, the analysis was exploratory, so no distribution test of the dynamic results was performed.

## RESULTS

The average clinical severity score of the patients was 3.05 ± 0.88 and the average length of the EcoRI fragment was 25.0 ± 5.9 KB0. The average results of the dynamometric evaluation were 28.6 ± 13.32 N and of the 6MWT 443.1 ± 138.3 m. The datasets of patients and HCs were tested for differences (see Table 1) and there was no indication that there were distribution differences in BMI, weight and height. However, there were some differences in age and in the distance of electrodes. Regarding the distance between the electrodes, there is a difference in the upper part of the distribution (max for FSHD: 24.5 cm, max for HCs: 18.5 cm). Regarding the age, the distribution for the HCs was slightly shifted to higher values in respect to patients.

**Table 1.**
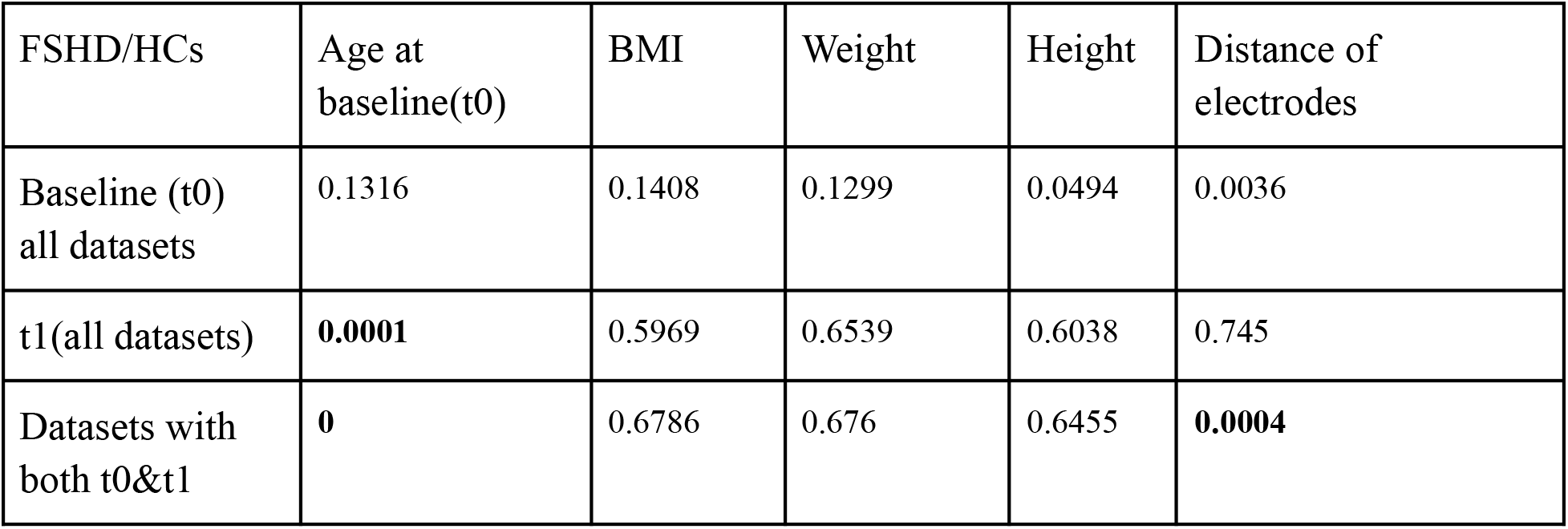
p-values from the t-tests of the comparison between FSHD patients and healthy controls (HCs). The level of significance after Bonferroni correction was 0.003. BMI: Body-Mass-Index.

Dynamic data could not be acquired for every scan due to various technical reasons that were unrelated to the patients’ clinical condition or to their response to the scan during stimulation. Therefore, in some cases, dynamic data were missing for an intermediate point or for one side (e.g. the dynamic scan was only performed on the left or right thigh). A total of 177 dynamic scans for FSHD subjects and 39 for HCs were acquired (see Table 2). An overview of the exact datasets included in this analysis is presented in Table 2. The time difference between various time points was not always precisely 6 months, but since differences were small and no rapid FSHD-caused change was expected, the analysis was performed as per time point.

**Table 2.** Overview of dynamic datasets included in the analysis.

|  |  | Time Points (6-month interval) | **t0**<br>**baseline** | **t1** | **t2** | **t3** |
| --- | --- | --- | --- | --- | --- | --- |
| FSHD Patients |  | Number of subjects | 34 | 24 | 22 | 12 |
|  | Single Side Datasets (ssd) | Nr. of ssd (177 in total) | 65 | 46 | 43 | 23 |
|  |  | Nr. of ssd with **min 2** time points | 56 | 46 | 43 | 23 |
|  |  | Nr. of ssd with **min 3** time points | 41 | 32 | 39 | 22 |
|  |  | Nr. of ssd with **only 1** time point | 9 | 0 | 0 | 0 |
|  |  | Nr. of ssd with **only 2** time points | 15 | 14 | 4 | 1 |
|  |  | Nr. of ssd with **only 3** time points | 30 | 21 | 28 | 11 |
|  |  | Nr. of ssd with **all 4** time points | 11 | 11 | 11 | 11 |
| Healthy Controls |  | Number of subjects | 12 | 7 | 2 |  |
|  | Single Side Datasets (ssd) | Nr. of ssd (39 in total) | 23 | 12 | 4 |  |
|  |  | Nr. of ssd with **min 2** time points | 11 | 12 | 4 |  |
|  |  | Nr. of ssd with 1 time point | 12 | 0 | 0 |  |
|  |  | Nr. of ssd with 2 time points | 8 | 9 | 1 |  |
|  |  | Nr. of ssd with 3 time points | 3 | 3 | 3 |  |

As explained above, the applied current for NMES was chosen in the first MRI session, but for subsequent visits it was often not possible to maintain the same current setting, mainly because of discomfort reported by the subject. As reported in Fig. 2, in the case of FSHD patients the applied current had to be only slightly modified between scans, whereas for HCs the current was often increased between the first and the second scan. Specifically, for FSHD patients, the applied current was generally slightly decreased in respect to the first scan, with the median current changes of -1.5 mA (*t1*), -2 mA (*t2*), and -4 mA (*t3*). For HCs the median increase of current between the first and the second scan was + 3mA (*T1*). The effective slope of the changes (in reference to the first scan) in current versus changes in strain was 0.00455 (intercept=-0.01125, p=0.000108). Therefore, a multiplication with 0.00455 was used for normalization.

**Figure 2.**
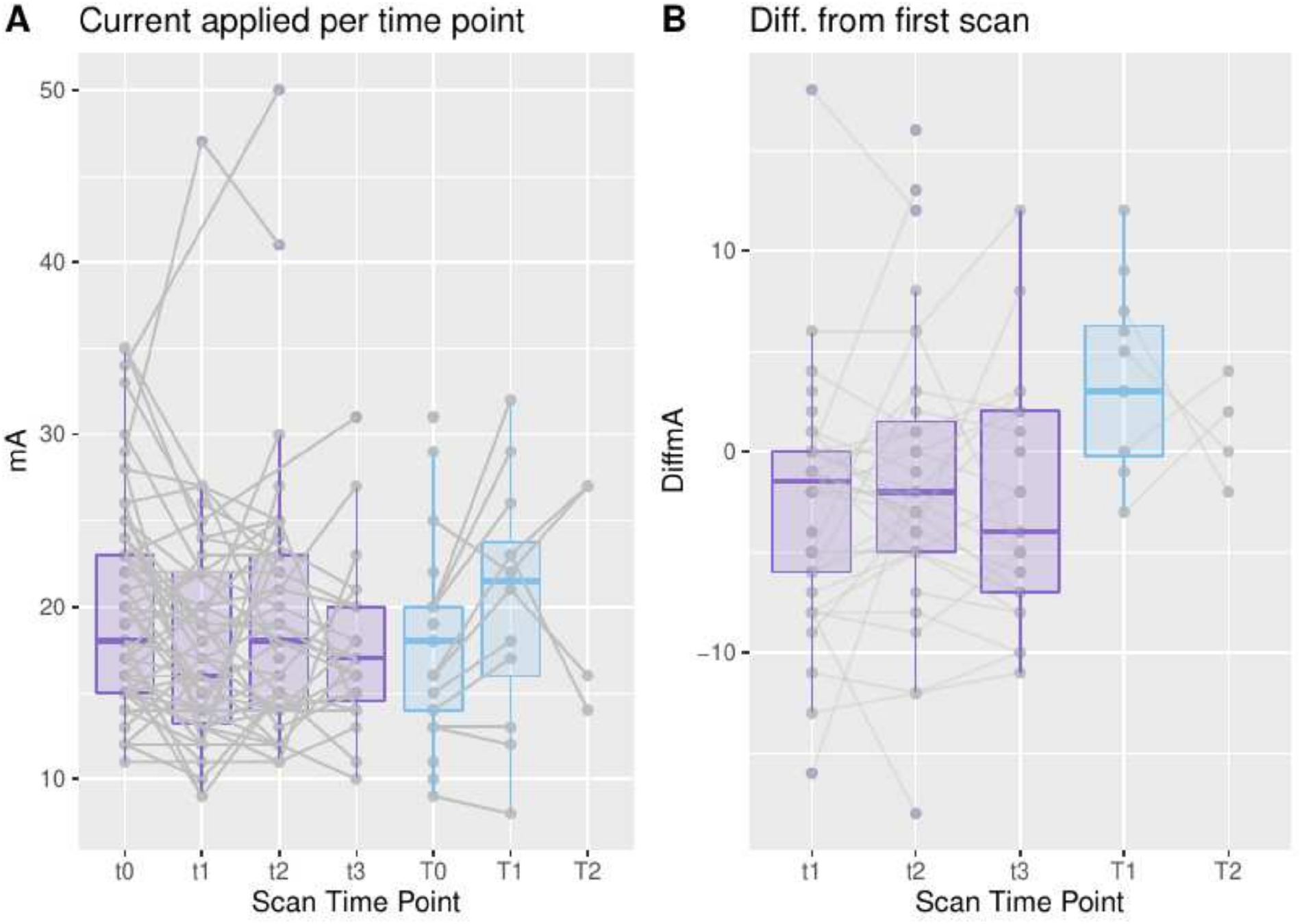
Visualization of the settings of current for every respective scan in absolute values as well as differences (in reference to the respective first scan). A) Current that was applied for the respective time point(*left*: t0-t3 for FSHD patients, *right*: T0-T2 for healthy controls), B) Differences in current in mA from the first respective scan calculated for every subject individually.

An overview of the dynamic results in the baseline (65 datasets for patients, 23 for HCs) is presented in Fig. 3, as well as the respective applied current. The strain values for the FSHD patients as a group were higher than for HCs, as was the applied current (see Fig. 3A&B). Similarly, the absolute values of both the build-up and release rates were higher for the FSHD patients than for the HCs. At the first follow-up time point (88 datasets for patients, 12 for HCs, see Fig 4), the strain of the FSHD patients as a group was lower than for the HCs, along with the current, while the rates for the FSHD patients were reduced in comparison to the volunteers.

**Figure 3.**
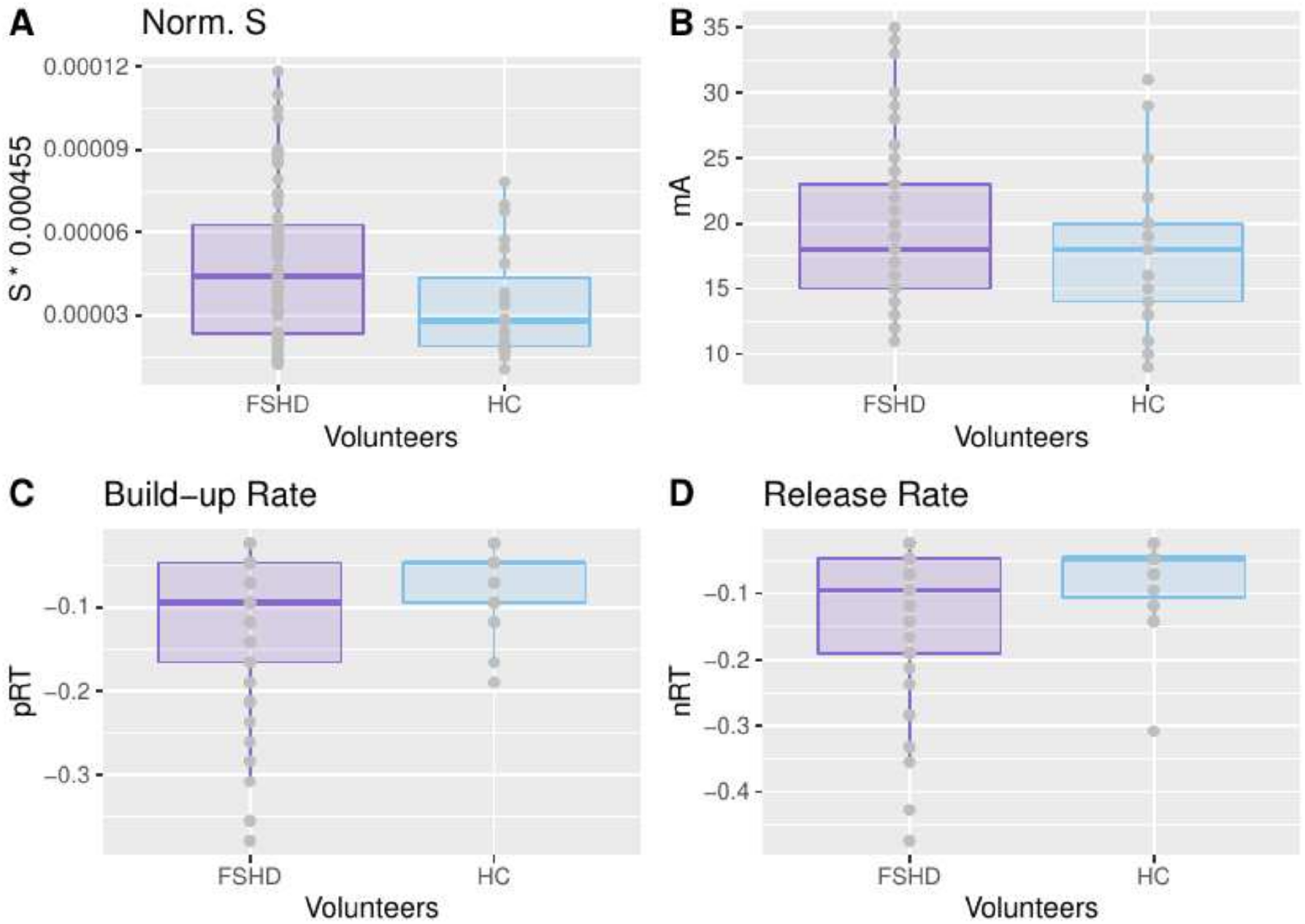
Dynamic results at baseline (time point 0, 65 for FSHD, 23 for HCs). A) Normalized strain at baseline for FSHD patients and healthy controls (HCs). B) Current applied in mA, C&D) Build-up and release-rates at baseline. Here, data is reported as a total with no differentiation.

**Figure 4.**
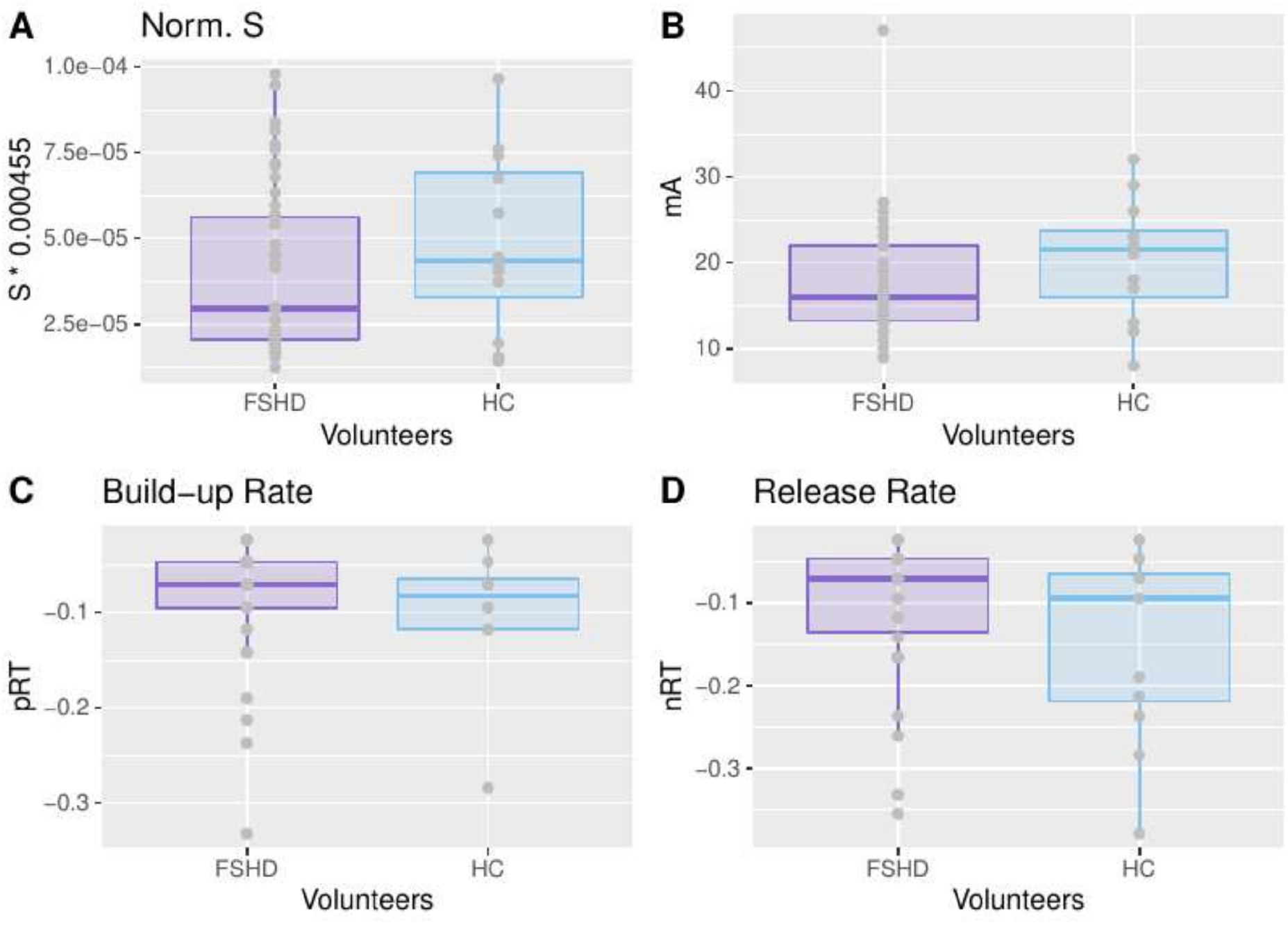
Dynamic results at time point 1 (88 datasets for patients, 12 for HCs). A) Normalized strain at baseline for FSHD patients and healthy controls (HCs). B) Current applied in mA, C&D) Build-up and release-rates at baseline. Here, data is reported as a total with no differentiation.

The comparison between patients and HCs was lastly performed for the groups with the same numbers of data points at the baseline and at time point 1 (see Table 2): 44 datasets for the FSHD patients (both t0 & t1) and 11 for the HCs (both T0 &T1). Fig. 5 depicts the respective normalized strain values. For FSHD patients the strain was decreased at the second time point, while for HCs it increased (see Fig. 5A). The range of absolute strain values did not differ between the two groups (Fig. 5). However, the median normalized difference over all datasets of the patients was close to 0 (6.825e-06, see Fig. 5A&D). Therefore, for clearer visualization, the results were differentiated based on an increase (see Fig. 5-B, E, H) or decrease (see Fig. 5-C, F, I) in strain with respect to the first scan. The first patient group (DiffS>0, Fig. 5B, E, 5) showed similar trends to the HCs. Moreover, the differences in strain between the patients and the HCs seem to be independent from the applied current (see Fig. 5H). The second patient group (DiffS<0, Fig. 5C, F, I) showed a decrease in strain. For the rest of the manuscript, the first group (DiffS(T1-T0)>=0) will be referred to as FSHD-Δs+ group, while the second (DiffS(T1-T0)<0) as FSHD-Δs-.

**Figure 5.**
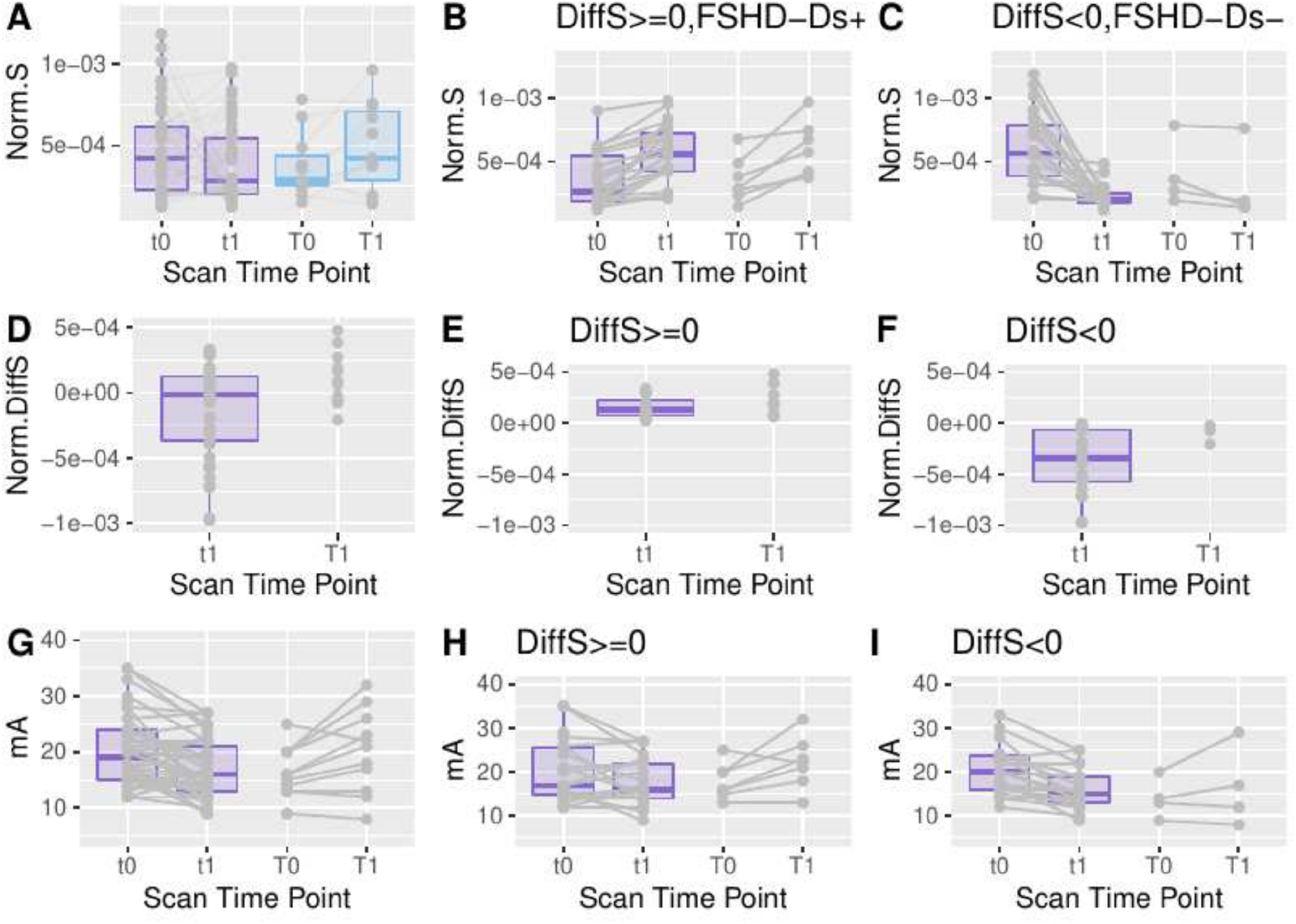
Comparison of FSHD patients and healthy controls (HC) for the first two time points for data with equal number of datasets for baseline and for time point 1. **A**) Normalized strain in all datasets without differentiation (*1st column*), **B**) Normalized strain in datasets that present increase of strain (22 for FSHD, 7 for HCs, *2nd column*), **C)** Normalized strain in datasets that present decrease of strain (22 for FSHD, 4 for HCs, *3rd column*), **D-F**) Normalized differences of strain, in relation to the first scan, for the respective datasets in A-C, **G-I)** Current values that were applied in the respective cases. The datasets in the 2nd column (DiffS(S1-S0)>=0, FSDH+) are very similar to the HCs, while the datasets in the 3rd column (DiffS(S1-S0)<0, FSHD-Δs-) present a decrease in strain different than in HCs.

The datasets of FSHD patients for the remaining time points were visualized separately from the HCs since more data points were available for this group (see Fig. 6). The single side datasets (i.e., one leg) that had a minimum of three out of four time points were selected since these datasets were the most abundant (see Table 2). The normalized strain for these datasets and the differences in strain with respect to the first scan were visualized, after distinguishing the FSHD-Δs+/FSHD-Δs-groups (Fig. 6). For the FSHD-Δs+ group there is no clear trend, whereas for the FSHD-Δs-group there is a decreasing trend which is more pronounced between the first two time points. The median of all datasets between the first and third time point was decreased by approximately 47%.

**Figure 6.**
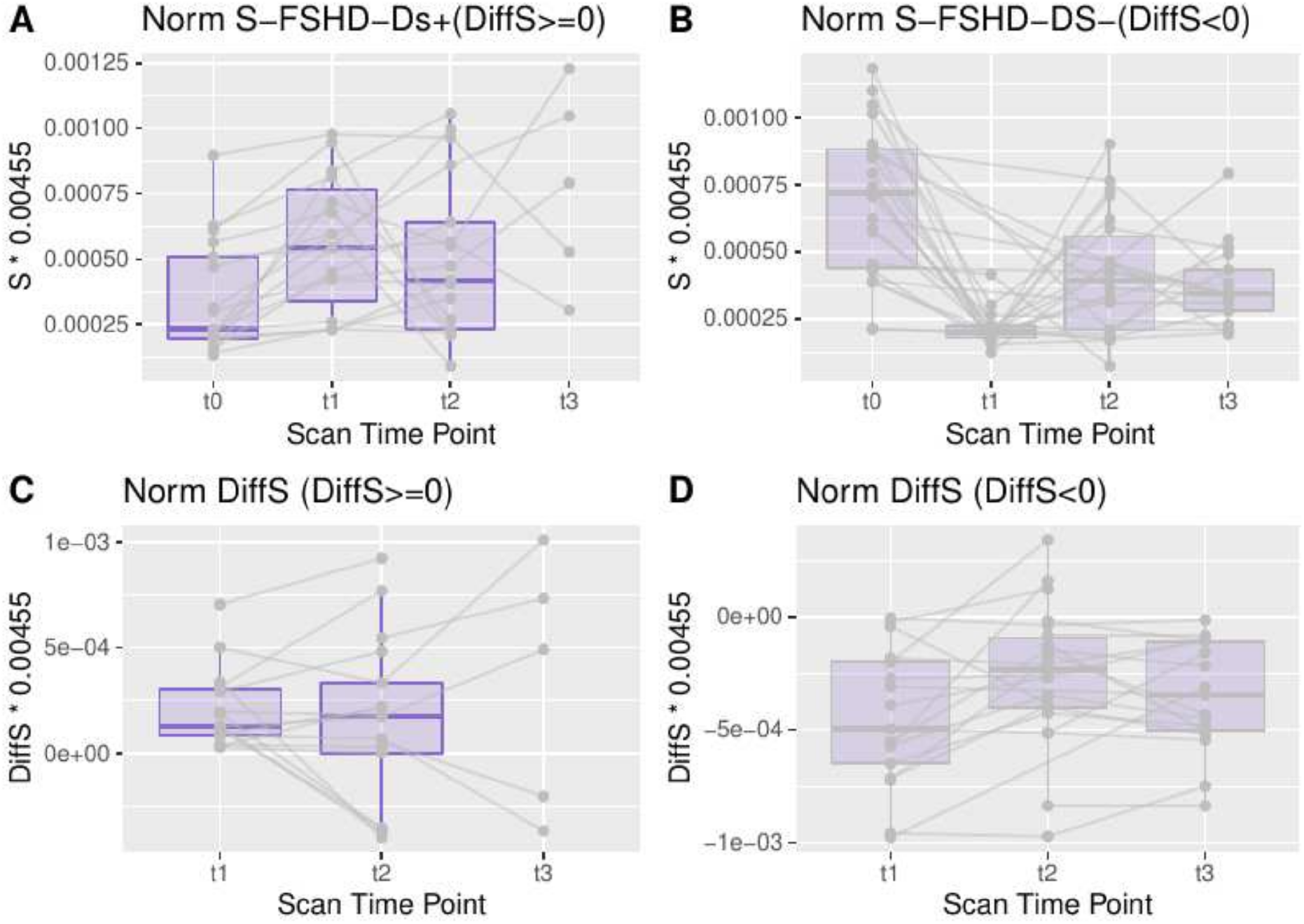
Datasets for FSHD patients, including a minimum of three time points (any combination out of the 4 time points). (A) normalized strain for the FSHD-Δs+ datasets (17 (**t0**), 15 (**t1**), 17 (**t2**), 5 (**t3**)) (see Fig.3). B) normalized strain for the FSHD-Δs-datasets (24 (**t0**), 17 (**t1**), 22 (**t2**), 17 (**t3**)) (see Fig. 3). Respective differences in the strain from the first time point. For the FSHD-Δs+ group (DiffS(S1-S0)>0), the median difference from the first time point remains stable (with an increasing variability). For the FSHD-Δs-group (DiffS(S1-S0)<0), there are larger fluctuations in the difference from the first time point and the absolute difference is larger between the first 2 time points than in subsequent ones.

The build-up and the release rates, similarly to the strain in the datasets where both the baseline and the first time point were available, were also analyzed separately. Larger absolute values indicate steeper time evolution (Fig. 7). For the FSHD-Δs+ group (Fig. 8A&C) the rates (in absolute value) were increased in the 2nd time point (t1) comparably to the HCs (T1). For the FSHD-Δs-group (Fig.8 B&D) the rates were decreased in the 2nd scan, a trend that was not observed in the HCs.

**Figure 7.**
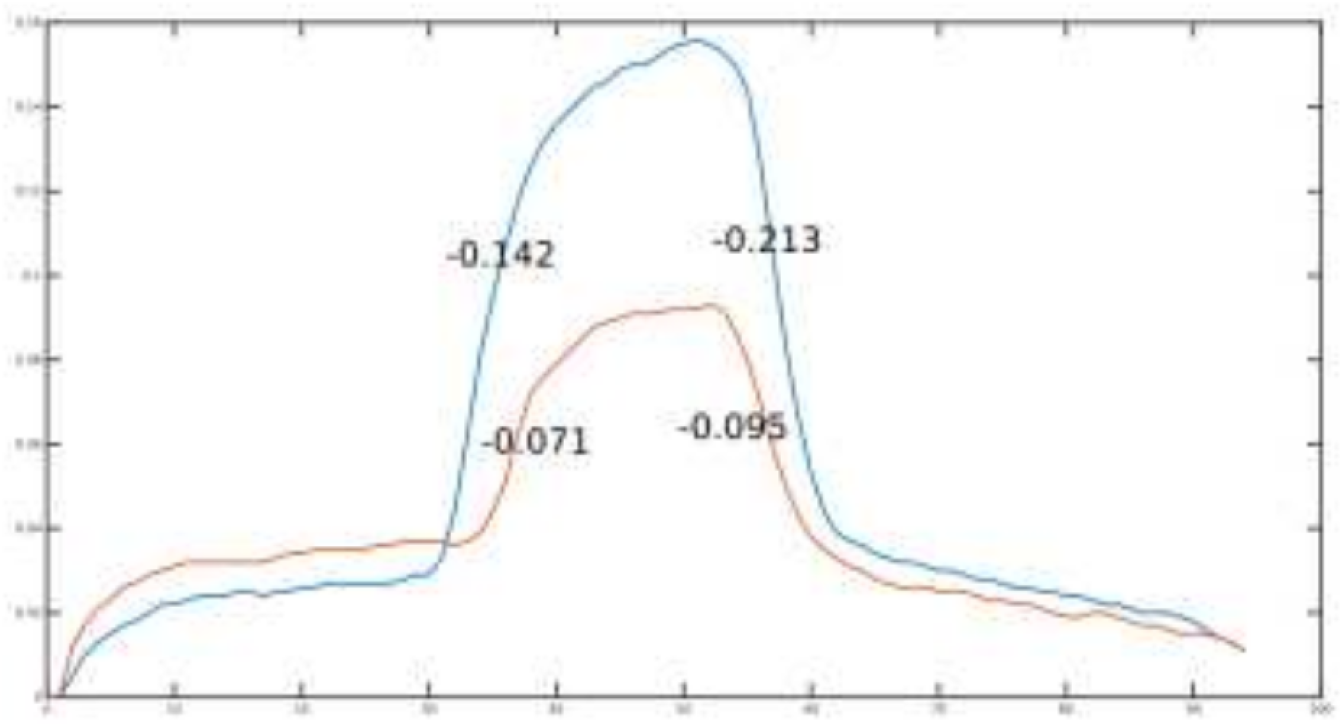
An example of strain curves and respective strain evolution versus phase. A steeper curve is expected to correspond to a more elastic muscle.

**Figure 8.**
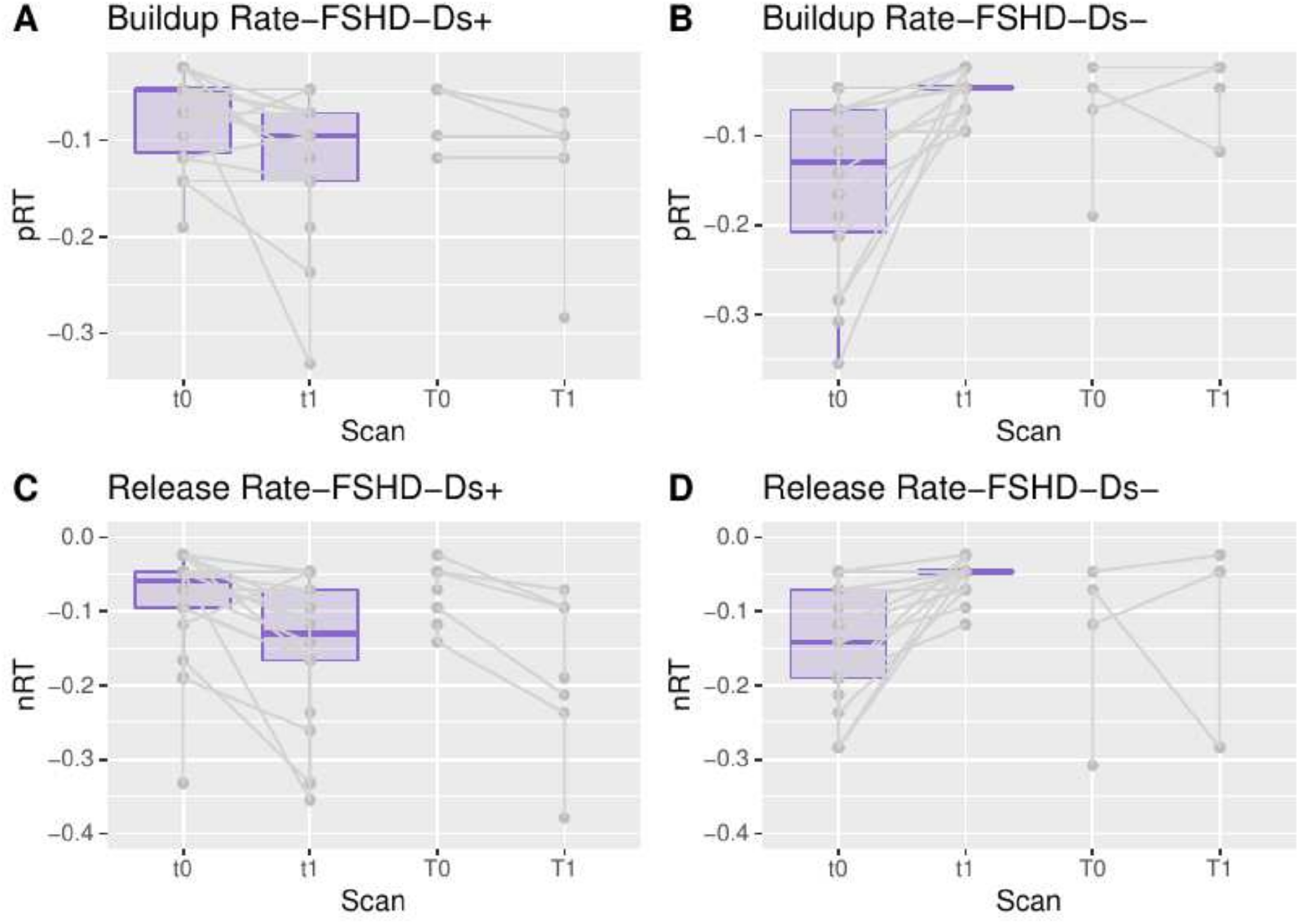
Build-up & release rates comparison between patients and healthy controls (HCs). **A**) Build-up rate for the first two scans for the FSHD-Δs+ patient group and the HC group (22 for FSHD, 7 for HCs), **B**) Build-up rate for the FSHD-Δs-patient group and the HC group (22 for FSHD, 4 for HCs), **C**) Release rate for the first two scans for the FSHD-Δs+ patient group and the HC group (22 for FSHD, 7 for HCs), D) Release rate for the FSHD-Δs-patient group and the HC group (22 for FSHD, 4 for HCs).

Lastly, the CSS, average length of the EcoRI fragment, 6MWT and dynamometry results were compared for the two groups (FSHD-Δs+/ FSHD-Δs-, see Fig. 9). The FSHD-Δs+ group had higher dynamometry results and KB0 values, whereas there was no difference between the two groups in CSS values or 6MWT results. In addition, the age at baseline and distance of electrodes were also visualized between the two groups since these were different in the initial population comparison, but no differences were observed.

**Figure 9.**
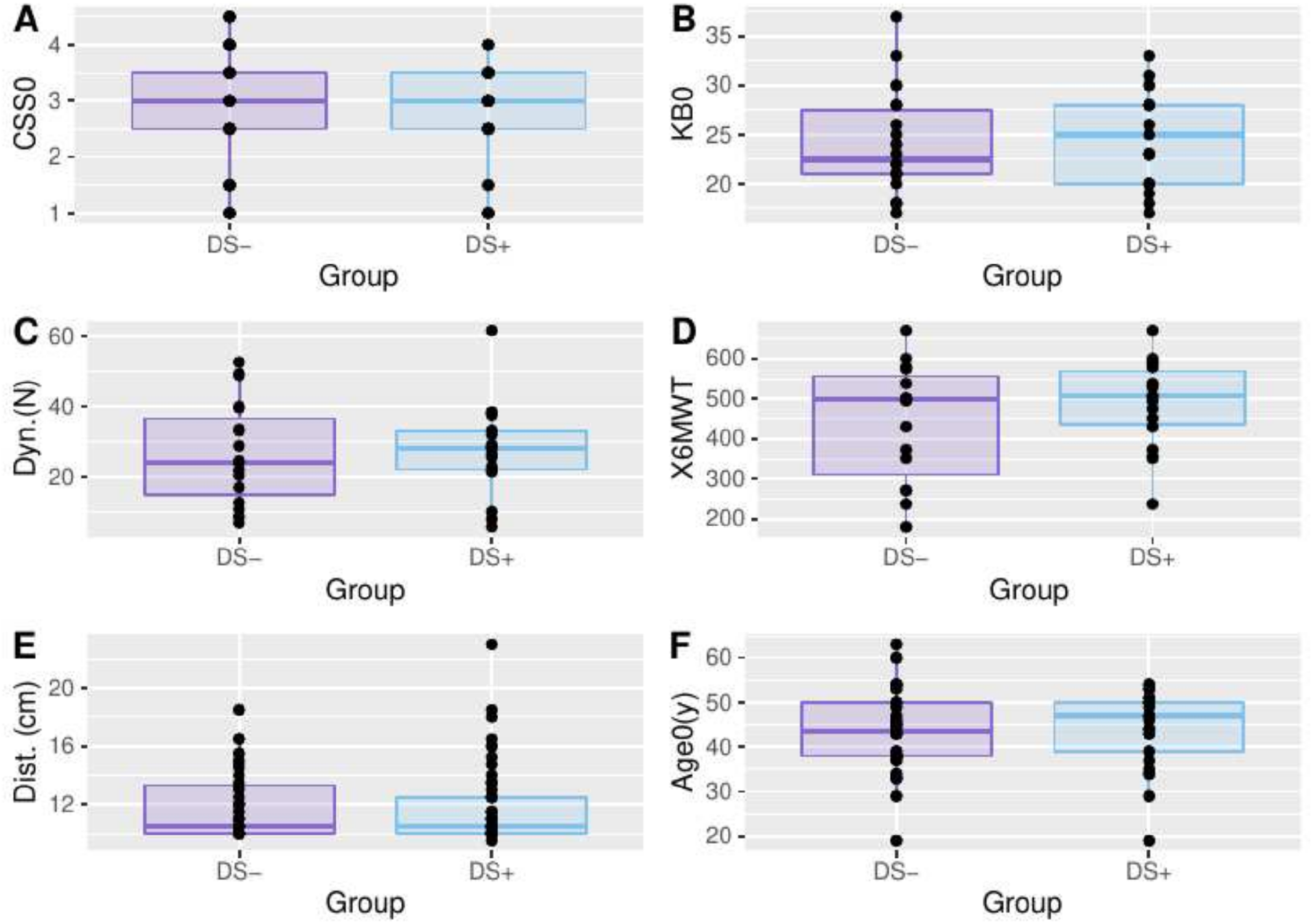
Functional results for the two patient groups FSHD-Δs+/FSHD-Δs-: A) Clinical Severity score at baseline (CSS0), B) Length of the 4q35 BlnI resistant, p13-E11 EcoRI fragment (KB0), C) Dynamometric evaluation (N), D) 6 minute walking test. In addition, the distance between the 2 electrodes (E) and the age at baseline (F) are given for the two groups.

Finally, water T2 and fat fractions values from all available time points were grouped into patient results versus HCs (FSHD: 25 datasets (t0), 17(t1), 9(t2) & HC (7(T0))). The mean values are presented on Table 3. In addition, the correlation coefficients of strain, buildup, and release rates versus the fat fractions and water T2 values of the vastus lateralis and vastus intermedius were calculated for all volunteer datasets together (Table 3). There were no significant correlations between max strain values and T2 or FF values (p>0.17). Similarly, regarding the rates (Table 3), min p-value for the calculation of the correlations was 0.083 indicating no significant correlation.

**Table 3.** Correlation coefficients (r) of the strain (S), build-up (pRT) and release rates (nRT) with water T2 (T2) and fat fraction (FF) in the Vastus Lateralis (VL) and Vastus Intermedius (VI). The significance level corrected for multiple comparisons was 0.0042.

|  | parameters | r | p | T2 (ms) or FF(%) in FSHD | T2 (ms) or FF(%) in HC |
| --- | --- | --- | --- | --- | --- |
| Strain | S_T2VL | 0.204 | 0.207 | T2VL: 43.13 ± 8.61 | T2VL: 39.44 ± 2.83 |
|  | S_T2VI | 0.215 | 0.176 | T2VI: 44.22 ± 7.89 | T2VI: 39.20 ± 1.95 |
|  | S_FFVL | 0.151 | 0.339 | FFVL: 14.59 ± 18.02 | FFVL: 6.39 ± 2.54 |
|  | S_FFVI | -0.161 | 0.309 | FFVI: 15.34 ± 18.94 | FFVI: 5.01 ± 2.06 |
|  | pRT_T2VL | -0.262 | 0.103 |  |  |
|  | pRT_T2VI | -0.256 | 0.107 |  |  |
| Build-up rate | pRT_FFVL | -0.184 | 0.243 |  |  |
|  | pRT_FFVI | 0.097 | 0.541 |  |  |
|  | nRT_T2VL | -0.236 | 0.143 |  |  |
|  | nRT_T2VI | -0.202 | 0.206 |  |  |
|  | nRT_FFVL | -0.127 | 0.423 |  |  |
| Release rate | nRT_FFVI | 0.27 | 0.083 |  |  |

## DISCUSSION

In this study, we explored the applicability and potential role of dynamic MRI in the field of neuromuscular diseases, applying it to a population of FSHD subjects and HCs, also evaluating the evolution of dynamic parameters over time. The acquisition protocol, with specific regard to the electrical stimulation during MRI scan, was well tolerated by FSHD subjects. Most existing similar MRI-studies focus on comparing younger and elderly healthy individuals (23,35,36) highlighting differences in muscle composition. To our knowledge, there is no study applying such a method to a cohort of patients affected by neuromuscular diseases. Here, the quantitative results of dynamic MRI of the quadriceps muscle contraction were different in the longitudinal evaluation for FSHD patients with respect to HCs. The range of strain values did not considerably differ between patients and controls, in contrast to what we would expect (23). Healthier muscles are expected to yield higher strain values indicating a better capacity of muscle deformation, while for instance atrophic muscle is expected to have lower strain values (37). However, according to their behavior in longitudinal measurements, there were two distinct groups of FSHD patients, one with similar strain changes (both magnitude and rates) to HCs (FSHD-Δs+) and another one with different trends (FSHD-Δs-), decrease in strain and also lower dynamometric results.

Here, in addition to the maximum strain, the build-up and release rates of the temporal evolution of the strain curve were also analyzed. The global range of rates was not significantly different between the groups of patients and volunteers, similarly to the maximum values of strain. From the comparison between HCs and FSHD patients for the first two time points, we observed two different groups of patients who showed either decrease (FSHD-Δs-) or increase (FSHD-Δs+) in strain in respect to the first scan. For patients with an increase in strain (FSHD-Δs+), the changes in the rates are very similar to the respective volunteer group. For patients with decrease in strain (FSHD-Δs-), the rates remain decreased across all the subsequent time points. The FSHD-Δs-group had similar CSS values and 6MWT results, but lower number of fragments and dynamometric results than the FSHD-Δs+ group. Due to the subdivision and strict selection of data, the two respective groups of HCs were rather small so further conclusions cannot be made at this point. The last point to mention is that the changes of build-up and release rates of the strain were less dependent on the current applied.

T2 and FF were not correlated to the absolute maximum strain value or rates. Although Andersen et al. (8) did not find a change of muscle strength in FSHD patients on quantitative MRI results, only FF was used in that study. Here, the dynamic results were also compared to T2 changes rather than just FF, but overall no significant correlation was observed as well. The results of the dynamic results presented here do not seem to coincide with standard quantitative MRI measures, leading us to hypothesize that they could provide additional information on skeletal muscle tissue composition other than relative fat or water content, and with particular regards to fibrosis. Due to the low number of datasets, further experiments are needed to investigate this hypothesis.

In general, for the group of HCs the current was increased in the second time point, which is probably due to exaggerated perceived discomfort in the first visit. Volunteers who are not used to regular medical procedures are more inclined to overreact in the first experiment with NMES, therefore not accepting the minimum optimal current for the experiment. In a second visit, being used to the complete procedure (NMES and MRI), it is more likely that they accepted an increased current. On the contrary, chronic patients have a higher resistance and acceptance limit of medical procedures, therefore as we see here the changes of current between time points were less pronounced (i.e., there is a fluctuation but not a trend). This finding suggests that the psychological state can be a factor in the muscle response to electrical stimulation, which would explain the slight increase in the strain values observed in the HC and FSHD-Δs+ groups. This potential effect could be evaluated with the addition of a perception questionnaire in the future.

A previous study investigating the reproducibility of MR imaging of NMES-evoked contractions during plantar flexion in healthy volunteers indeed showed a similar trend when the force output was kept constant (22). However, in comparison to previous studies with young healthy volunteers (38) the increased necessary current is unexpected, but here the age group and the time interval between experiments are very different. Finally, there was no obvious dependence of the build-up and release rates to the applied current.

There were a few limitations in this preliminary study. Here, the investigated muscles were not chosen based on the patients’ condition (i.e., most severely affected muscles to be investigated). Instead, the quadriceps muscle group was chosen for the sake of robustness, simplicity, and replicability due to its size, shape, and location (21,23). Also, the strain values analyzed were the maximum over a broader region of interest including both VL and VI muscles and a separate analysis of strain in VL and VI was not performed. Moreover, the number of participants was relatively low, and the number of HCs was lower than the number of FSHD patients. However, this was a pilot study and the subjects were scanned in exactly the same conditions and repeatedly.

In addition, in this study, there was no measurement of the force evoked by the NMES in order to standardize the stimulus, since this would require additional MR-compatible equipment. Therefore, no force measurements were included and we designed the protocol to induce a submaximal isometric contraction defined as the “minimum visible muscle twitch,” so no measurable force output would be expected (i.e., in line with (23)). In addition, force measurement is particularly complicated in the case of electrical stimulation of the thigh, because the response of knee extension would ideally have to be measured inside the bore of the MR scanner. To ensure reproducibility over time, the position of the electrodes was fixed by measuring the exact position of the electrodes at every visit. When possible, the current of the stimulation remained the same, under the condition however that a contraction was induced and the volunteer was not in discomfort. Nevertheless, the current in various cases had to be modified and a bias due to this cannot be completely excluded when evaluating strain, but the normalized changes in strain were also calculated and were expected to be independent from changes in the current.

Our results suggest that evaluating the muscle ability to contract and deform could offer useful insight to the evolution of FSHD and potentially other myopathies on mostly superficial muscles, at least in the longitudinal follow-up of the disease. It should be noted that the current setup was mainly dictated by the technical possibilities available when the study was initiated; recent advances in the acquisition method now enable three-dimensional coverage of the muscles (22,39) and might offer higher flexibility and diagnostic value, thus warranting further investigation.

## CONCLUSION

In conclusion, the quantitative results of strain MRI through evoked NMES could offer additional information to a standard quantitative MRI protocol, possibly highlighting subtle changes before they become detectable with other quantitative MRI biomarkers. More extensive studies are needed to support the findings of the current study that shows a promising direction potentially applicable to other neuromuscular disorders.

## Data Availability

Patient data are available upon request and after institutional agreement to be addressed to Anna Pichiecchio

## DECLARATIONS

### Ethics approval and consent to participate

The study was performed in compliance with the local IRB regulations (at the Mondino Foundation by the “Comitato Etico Area Referente Pavia Fondazione IRCCS Policlinico San Matteo”-P-20200042370 and the Policlinico Gemelli’s Ethics Committee-Prot. 7451/18 ID 1952.) and all volunteers signed an informed participation agreement.

### Consent for publication

Not applicable. No identifiable data are included.

### Availability of data and materials

Raw data are available upon request.

### Authors’ contributions

- **XD**: analysis, interpretation of data; drafted the manuscript, design of the work
- **FSa**: data analysis, substantively revised the manuscript, design of the work
- **MP**: Data acquisition, substantively revised the manuscript, design of the work
- **FSo**: Data acquisition, analysis,
- **NB**: Data acquisition, analysis, substantively revised the manuscript
- **GS**: Data acquisition, substantively revised the manuscript
- **AF**: Data acquisition, **GG**: Data acquisition,
- **MM**: conception/design of the work, substantively revised the manuscript
- **GT**: conception/design of the work, substantively revised the manuscript
- **ER**: conception/design of the work, substantively revised the manuscript
- **AP**: Data acquisition, conception/design of the work, substantively revised the manuscript

## Acknowledgements

- Not applicable

